# Challenges and facilitators in pathways to cancer diagnosis in Southern Africa: A qualitative study

**DOI:** 10.1101/2025.01.14.25320521

**Authors:** S. Day, KD. Arendse, SE. Scott, M. Moyo, S. Mzeche, BT. Guzha, N. Tegama, VA. Sills, T. Ras, FM. Walter, J. Moodley

## Abstract

**Objectives:** To explore healthcare workers (HCW) experiences, barriers and facilitators in managing patients with symptoms of possible breast, cervical or colorectal cancer.

**Design:** A qualitative in-depth interview study with HCWs managing patients with breast, cervical and colorectal cancer symptoms. We also conducted workshops with a group of HCWs to check the credibility of the interview findings.

**Setting:** The study was conducted with staff working in primary, secondary and tertiary health facilities in the Eastern and Western Cape in South Africa (SA) and Harare and Bulawayo and their referral provinces in Zimbabwe.

**Participants:** HCWs with experience in managing patients with symptoms of possible breast, cervical or colorectal cancer were recruited for the study. Participants were purposively sampled, according to region, healthcare level, and job role. A total of 56 participants (26 in SA and 30 in Zimbabwe) participated in the in-depth interviews. Twenty-six (12 in SA and 14 in Zimbabwe) participated in four clinical advisory group workshops across both countries.

**Results:** Drawing on the Model of Pathways to Treatment, HCW perceptions of patient-level factors influencing the diagnostic interval included financial limitations, and patients’ absence and delays in attendance. Healthcare provider and system factors included: challenges with referral and feedback systems; training needs; low awareness of protocols and guidelines; inappropriate and suboptimal clinical assessments; and broader socio-economic factors and resource limitations.

**Conclusion:** Improving the timely diagnosis of breast, cervical, and colorectal cancer in Southern Africa necessitates targeted strategies that address both patient-related, provider and health-system delays.

**Strengths and Limitations:**

- To our knowledge, this is the first study in SA and Zimbabwe that explores healthcare workers across all levels of care perceptions of the challenges and facilitators in the pathways to breast, cervical and colorectal cancer diagnosis.
- The qualitative nature of the inquiry and strong theoretical underpinning with the Model of Pathways to Treatment enabled in-depth exploration of the research question.
- The study setting included both urban and rural settings that represent differences in access to cancer care and health systems.
- We note that the discussion of patient level facilitators and barriers to early diagnosis are the perceptions of HCWs and not the views of the patients themselves.
- By examining cancer care in two distinct countries, the study provides valuable insights into how different levels of human development index impact healthcare systems and identify unique challenges and best practices. A comparative analysis reveals important differences in health outcomes and resource allocation, which can inform targeted interventions.

## Introduction

Globally, cancer is a leading cause of death accounting for nearly 10 million deaths in 2022, with a predicted increase to 18.5 million cancer-related deaths per year by 2050, of which 73% will occur in Low- and middle-income countries (LMICs).^1^ The incidence of cancer is predicted to rise from 20 million in 2022 to 35.3 million per year by 2050.^1, 2^ LMICs face a disproportionate burden of cancer-related morbidity and mortality.^3^ Increasing cancer incidence and poorer prognosis in LMICs can be attributed to ageing societies, health disparities, limited access to quality healthcare, and high prevalence of risk factors.^3^ High proportions of patients in LMICs are diagnosed with advanced-stage cancer^4–6^ due to delayed presentation, poor referral pathways and limited diagnostic capacity.^7, 8^ Global disparities in cancer mortality and survival rates are marked by differences in clinical stages at diagnosis.^9^ The World Health Organisation (WHO)^10^ Report on Cancer: ‘Setting priorities, investing wisely and providing care for all’ recommends prioritising early diagnosis programmes, including screening and timely diagnosis of symptomatic people, with access to quality treatment in the absence of universal, equitable screening programmes.

Considering the burden of breast, cervical and colorectal cancer in Southern Africa,^11^ strategies and interventions are needed to ensure timely diagnosis. Improving earlier diagnosis of cancer in Southern Africa requires understanding pathways from symptom recognition to diagnosis^6, 12^ where both patient-related and health system-related delays are common.^6, 13^ The WHO 2020 Report on cancer identifies key events across the cancer continuum and conceptualises them as three phases: from symptom awareness (phase 1) to diagnosis (phase 2) and treatment (phase 3).^14^ The Model of Pathways to Treatment^15^ also identifies the interval phases and underpinning patient-, disease-, health system- and provider-related factors that contribute to time to diagnosis and treatment.^4, 6, 13, 15–17^ The focus of this study is the ‘Diagnostic Interval’, defined in the Model of Pathways to Treatment as the time between the first encounter with a healthcare worker (HCW) and the formal diagnosis being made. Improving efficiency in the diagnostic interval has the potential to increase early detection,^18, 19^ and facilitate downstaging.^20^ The Diagnostic interval is complex and involves initial clinical assessment, usually by primary-level HCWs, and subsequent investigations in order to triage and make a judgement about the diagnosis. The diagnostic process requires available diagnostic resources and staff alongside effective referral and feedback mechanisms between primary, secondary and tertiary facilities.^21–23^

There has been relatively little exploration of the Diagnostic interval for cancer diagnosis in Southern Africa, although exploratory work has suggested that context-relevant training for primary-level HCWs, improved referral pathways and additional human resources may be required to reduce the Diagnostic interval.^12, 13, 24, 25^ Moodley et al.^26^ have demonstrated that primary-level HCW clinical assessment of breast and cervical cancer in South Africa (SA) is influenced by competing health priorities, such as HIV, infectious diseases, violence and injury, and maternal and child health, and other non-communicable diseases. Patients with breast and cervical symptoms may return to the clinic several times before being referred due to cancer symptoms being mistaken for other common health concerns. A systematic review on routes to diagnosis of symptomatic cancer in Southern Africa reported no studies on breast, cervical or colorectal cancer in SA or Zimbabwe.^27^

This study therefore aimed to explore primary, secondary and tertiary level HCWs experiences, barriers and facilitators in managing patients with symptoms of possible breast, cervical or colorectal cancer in two Southern African countries with varying health system challenges: SA (in the medium human development index (HDI) category (0.709))^28^ and Zimbabwe (in the low HDI category (0.550)).^29^

## Methods

### Design

A qualitative in-depth interview study with HCWs managing patients with breast, cervical and colorectal cancer symptoms was undertaken. We also conducted workshops with a different group of HCWs to check the credibility of the findings from the interview findings by providing a second source of input and comparison for the interview findings. The project was part of the African aWAreness of CANcer & Early Diagnosis programme (AWACAN-ED) (awacan.online)^30^, aiming to advance cancer awareness and early diagnosis in Southern Africa through understanding the context of early cancer diagnosis in the region and developing culturally appropriate toolkits to improve timeliness of cancer diagnosis, with a specific focus on breast, cervical and colorectal cancers.

### Study setting

The study was conducted with HCWs working in public health facilities in the Eastern and Western Cape in SA and Harare and Bulawayo and their referral provinces in Zimbabwe. In the health sector in both countries, private and public healthcare systems run in parallel. The private sector – which services a smaller proportion of the population – is funded through individual contributions to medical aid schemes or out of pocket payments for those who do not have medical aid.^31^ The high costs of private healthcare and medical aid contributions places private healthcare out of the reach of much of the population.^32^ The public healthcare sector is made up of three tiers: primary, secondary and tertiary. Guidelines state that primary healthcare (PHC) clinics and community health centres should be patients’ first point of contact for most healthcare requirements and are the main mechanism through which health programmes are delivered. However, some people enter directly into secondary or tertiary facilities or have an emergency admission. In some cases, private general practitioners (GPs) may be the first point of contact for the health system for those who have personal financial means or medical aid/insurance. There is a hierarchy of hospital levels offering specialist and sub-specialist services, which make up secondary and tertiary facilities. Depending on the health facility resources, such as colposcopy, colonoscopy, imaging facilities, and the availability of specialist surgeons, patients will either be diagnosed and treated at the secondary facility, or they will be referred to a tertiary facility for diagnostic investigations and treatment.^33, 34^ In SA, PHC facilities are free to the public, while secondary and tertiary facilities use a Uniform Patient Fee Schedule – a means test – which sets out who should pay and how much they should pay to access healthcare.^35^ In Zimbabwe, patients pay fees at each healthcare level for consultations, medications, and procedures.^36–38^ In some rural areas in Zimbabwe, PHC facilities are free and some patients are exempt from fees, such as children under 5, people older than 65, people diagnosed with psychiatric disorders, people with disabilities, and pregnant women.

In SA, the policy for breast cancer screening involves opportunistic clinical examinations for all women when they present to primary care facilities for other health needs.^39^ Guidelines recommend that symptomatic patients receive immediate referrals to specialised breast units, not waiting more than 62 days for low-risk patients, and 21 days for high or medium suspicion patients for appointments.^39^ For cervical cancer, the policy recommends that symptomatic individuals should be referred immediately, although specific timelines are not established.^40^ Colorectal cancer lacks defined referral guidelines, but symptomatic patients should be promptly referred according to national cancer guidelines.^41^

In Zimbabwe, there are no specific management protocols for symptomatic patients.^42^ Cervical cancer guidelines recommend that patient with symptoms must be referred immediately, but the policy does not provide clear timelines.^43^ Like SA, colorectal cancer lacks established referral guidelines.

### Sample

Potential participants were eligible for inclusion if they were staff with experience of working with patients who presented with symptoms of possible breast, cervical or colorectal cancer. We used a matrix to purposively sample participants across the four regions, according to the level of their region, healthcare level, and job role.

### Procedure

Facility operation managers (primary care) or heads of department (secondary or tertiary) facilitated identification of potential participants for the in-depth interviews. Participants would sometimes recommend other potential participants.

Those who expressed interest after receiving a study information sheet were asked to provide written consent prior to interviews. The interviews were conducted by experienced qualitative researchers. Most interviews took place in a private setting at the health facility while two took place virtually, and all lasted between 30 and 90 minutes. Interviews were conducted in English or local languages (isiXhosa in SA, and Ndebele or Shona in Zimbabwe), based on participant preferences. In one interview, there were three additional HCWs present in addition to the main interviewee; questions were directed to and answered by the main interviewee, while the additional HCWs added details for context and clarity. Interviews conducted in a language other than English were translated prior to transcription. Interviews were audio-recorded and transcribed verbatim using a professional transcription company. At the start of each interview, socio-demographic details (age, job role, gender, education level and years of experience) were collected. A semi-structured approach was used following a topic guide which aimed to explore (i) barriers and facilitators in the pathways to timely cancer diagnosis and (ii) participants’ experiences and perceptions of the barriers and facilitators to digital technology use to support early cancer diagnosis in their daily practice (See the interview guide: supplementary file 1). This study focuses on the analysis of part (i) of the interview. Part (ii) is reported separately.^44^ Development of the first part of the interview schedule was guided by the Model of Pathways to Treatment^15^ and explored information related to current practices and HCWs experience of factors that influenced timely diagnosis of cancer. The interview guide included information specific to current practice and decision making such as how participants currently go about executing their roles and making decisions on next steps for symptomatic patients who present at facilities including information on perceived barriers and facilitators to timely cancer diagnosis.

Four workshops (two in each country) were conducted with clinical advisory groups to present interim findings and inform the development of interventions to promote timely diagnosis of cancer. These clinical advisory groups were made up of PHC practitioners and breast, cervical and colorectal cancer specialists at secondary and tertiary facilities. The workshops were structured around two parts. In the first part, we presented the preliminary results an ongoing AWACAN-ED cross-sectional, health facility assessment and the in-depth interviews. After, a facilitator led a discussion focused on the following areas: (i) key challenges for each type of cancer at the primary, secondary, and tertiary healthcare levels; (ii) current best practices for managing each cancer across these healthcare levels; and (iii) the characteristics of an ideal referral tool. This paper drew on information about key challenges and current best practices. Each workshop was audio recorded and transcribed. Notes were also taken in each workshop.

### Data Analysis

Codebook Thematic analysis was used to synthesise the interview data, starting with data familiarisation and coding.^45^ Coding is an iterative process whereby researchers seek to identify “a word or short phrase that captures and signals what is going on in a piece of data in a way that links it to some more general analysis issue”.^46^ The data had three coders. SD read and coded all the transcripts, and KDA and MM each coded 24 transcripts. Codes were collaboratively developed through an iterative process. After familiarisation with the data, we created a codebook describing each code with an example extrapolated from the data. We had code comparison discussions amongst coders, and disagreements and refinements on codes were resolved through discussion. After this process, themes were developed. Themes were used to attach significance to our findings, offer explanations, and draw conclusions. Through discussion, SD sense checked the analysis write-up with co-authors in Zimbabwe (SM, MM and BG) and co-authors with medical training (KDA, JM, TR, FW).

The transcripts and notes of the clinical advisory workshops were analysed thematically by VS to categorise the content of the discussions. This was then compared to the interview findings. Quotes in text are all from the interviews.

### Ethics

The study protocol received ethical approval from (i) The University of Cape Town, Faculty of Health Sciences Human Research Ethics Committee (HREC Ref: 664/2021 and HREC 892/2023), (ii) The Joint Research Ethics Committee (JREC) at The University of Zimbabwe (JREC Ref: 363/2021) and (iii) the Medical Research Council Zimbabwe (HREC Ref: MRCZ/A/2831).

## Results

For the in-depth interviews, 26 participants in SA and 30 participants in Zimbabwe were recruited, a total of 56 participants across both countries (see Table 1). In-depth interviews were conducted between July and November 2023. Less than half of the participants worked in primary care (n=24, 43%) and the remaining in secondary or tertiary care (n=32, 57%).

**Table 1:** Interview participant distribution across health facilities.

| Province | Primary Facilities | Secondary and Tertiary Facilities | Total |
| --- | --- | --- | --- |
| SA |  |  |  |
| Western Cape | 5 | 8 | 13 |
| Eastern Cape | 7 | 6 | 13 |
| *Zimbabwe and greater regions/ provinces* |  |  |  |
| Harare | 9 | 9 | 18 |
| Bulawayo | 3 | 9 | 12 |
| Total | 24 | 32 | 56 |

The participants’ median age was 44 years old (inter-quartile range: 34 - 53 years). Most of the participants identified as female (38/56). Median years in the role was 6 years (inter-quartile range: 2 – 15 years). Thirty-six (64%) participants’ highest education degree was an undergraduate degree or diploma, 16 (29%) had a postgraduate degree, 3 (5%) had Matric / O Levels and 1 (2%) had a certificate. Thirty-four (61%) nurses (20 primary/14 secondary & tertiary), 15 (27%) doctors (2 Primary/13 secondary & tertiary), 3 (5%) medical managers, 1 (2%) health information office, 1 (2%) clinical assistant, 1 (2%) data clerk and 1 (2%) receptionist were interviewed. Twenty-three (41%) participants managed patients with breast, cervical, and colorectal symptoms; 12 participants (21%) managed patients with breast and cervical cancer symptoms; 7 (13%) managed patients with cervical cancer symptoms; 6 (11%) participants managed patients with breast cancer symptoms; 5 (9%) managed colorectal cancer symptoms, and 3 (%) managed breast and colorectal cancer symptoms.

A total of 26 participants (12 in SA and 14 in Zimbabwe) (Table 2) participated in the clinical advisory workshops across both countries (see supplementary file 2 for more details).

**Table 2:** Clinical workshop overview.

| **Country** | **SA** |  | **Zimbabwe** |  |
| --- | --- | --- | --- | --- |
| **Location** | Cape Town | Cape Town | Harare | Bulawayo |
| **Number of clinicians** | 6 | 6 | 7 | 7 |
| **Clinician summary** | PHC and Specialists | Specialists | PHC and Specialists | PHC and Specialists |
| **Cancers covered** | Breast and cervical | Colorectal | Breast, Cervical and Colorectal | Breast, Cervical and Colorectal |

### Overview of findings

HCWs perceptions of patient-level factors influencing the diagnostic interval included (1) financial limitations impact on pathways to diagnosis, and (2) patients’ refusal, absence and delays in attendance. Healthcare provider and system factors included: (1) poor referral and feedback systems; (2) training needs; (3) awareness of protocols and guidelines; (4) inappropriate and suboptimal clinical assessments; and (5) broader socio-economic factors and resource limitations. Workshop findings are integrated into these themes and more details can be found in supplementary file 2. Quotes in the text are all from the in-depth interviews. The coders proposed quotes for inclusion, and the team reached a consensus to select those that best captured the themes’ meaning. Whenever possible, quotes were chosen from a diverse range of participants, representing different regions, healthcare levels, and job roles, to highlight both commonalities and contrasts among participants.

### HCWs perceptions of patient-level factors that impact the Diagnostic interval

#### Financial limitations impact on pathways to diagnosis

HCWs in both SA and Zimbabwe reported that patients’ resources impacted the journey to diagnosis. Particularly in Zimbabwe, HCWs in the in-depth interviews and clinical advisory workshops reported that patients lacked the necessary funds for initial and follow-up healthcare visits, diagnostic tests, and investigations for staging. Financial constraints may cause delays in investigations or bar patients from accessing health services altogether. In the SA clinical advisory workshops, HCWs reported that some patients may enter the healthcare system by visiting a private GP and then access public healthcare due to lack of funds to stay in the private sector.

> *“… some of them they can come to us but not having money for the lab investigations. So, it’s very difficult for the doctors to go ahead and do the necessary things for these patients. Some of them they lack funds to do the biopsy and some of them they don’t have funds for even simple lab investigations.” Nurse, Tertiary, Harare*

> *“The problem is the onus is on the patient or the client. We will refer the clients to the hospital for investigations to be done there. So, it’s upon the patient on when they will go there. Some will tell you, “We don’t have money”, and they will not be having that money to go to the hospital. Maybe when they get there, money is also needed to run those samples.” Nurse, Primary, Bulawayo*

In SA and Zimbabwe, HCWs reported in both in-depth interviews and the clinical advisory workshops that patients often lacked finance for transport to health facilities especially in rural areas which can delay further investigations and diagnosis.

> *“The problem is the issue of money for the patients to move from the health facility to the hospital. They have some challenges of money, even for bills to pay there they don’t have. So, they fear even to move from their homes to the district hospital saying, “We don’t have money to pay. We don’t have money for transport”.” Nurse, Primary, Harare*

> *“And sometimes you send the client to the hospital, and they don’t go to the hospital, because they don’t have money to get there.” Nurse, Primary, Eastern Cape*

#### Patients’ refusal, absence and delays in attendance

HCWs in both in-depth interviews and the clinical advisory workshops reported patient-level refusal of care and missed appointments were barriers to early diagnosis. Particularly with cervical and colorectal cancer, HCWs in both in-depth interviews and the clinical advisory workshops reported that patients sometimes decline examination and further testing, such as colonoscopies, because of fears of discomfort and pain, and invasive testing. HCWs felt broader societal (mis)understandings of what happens during these tests serves to exaggerate and misinform, resulting in reluctance to seek care. The HCWs in the clinical advisory workshops further elaborated and stated that there is insufficient patient trust in HCWs and the healthcare system which results in refusal of diagnostic investigations. This demonstrates a need for broader health education around investigations for cancer symptoms.

> *“There’s this whole conception of how unpleasant a colonoscopy is. And each person likes to exaggerate it more than the next person, and things like that. So, people delay having colonoscopies. They don’t want it because of the unpleasantness.” Nurse, Tertiary, Western Cape*

HCWs reported that patients move between provinces (SA) and countries (Zimbabwe) which results in missed appointments and subsequent referral, investigation and diagnostic delays. Patients missing appointments is therefore embedded in a broader sociopolitical landscape and economic climate – a history of segregation, land dispossession, and a migrant labour system – that results in transient populations moving between provinces in South Africa or from Zimbabwe to South Africa.

> *“[The] Migrant labour system [makes it difficult for patients to arrive on a dedicated day]. Because you will think you have your patient, the patient is in Cape Town. Job opportunities. There’s a season for harvesting grapes in Cape Town and all that. And then there will be that. They won’t come on their appointment dates.” Nurse, Primary, Eastern Cape*

> *“And also, the commitment must also come from the patient side, because, sometimes, they have that appointment, they know exactly when they’re supposed to go to Somerset, then they end up in Eastern Cape. And then they say, no, I was in Eastern Cape. Now, it is also your problem to make another appointment.” Nurse, Primary, Western Cape*

HCWs in both in-depth interviews and clinical advisory workshops reported that patients were difficult to follow-up after missed appointments due to giving incorrect phone numbers, changing phone numbers, not answering phone calls or reading text messages, losing cell phones or providing incorrect addresses.

> *“It is when the phone does not work, or the contact number does not go through because they sometimes give us numbers that are not working, and they go straight to voicemail. And then now you must look for a community worker for that place so now it becomes a long process.” Nurse, Primary, Western Cape*

> *“You will find someone has no address, has no contact details. So, if that client is due for LEEP [Loop electrosurgical excision procedure], maybe we will manage to source funds from the donor that we can do a camp. We try to follow those clients. It’s difficult to follow them because someone has no contact details.” Nurse, Primary, Harare*

In SA, community healthcare workers (CHWs), members of the community that serve and respond to health needs of the community^47^, assist PHC practitioners to follow-up with patients who are not contactable, which forms an important safety netting mechanism by facilitating better pathways to diagnosis by following up with patients who have abnormal Pap smear results and have missed appointments. However, tracing patients can take a long time, resulting in referral delays. Tertiary level HCWs reported that they were not aware of CHWs being used at PHC to trace patients who have missed appointments. A doctor in the Western Cape, SA reported that they have previously used the police to trace patients with biopsy results that indicate a cancer diagnosis.

> *“What works is the fact that while you’re doing this examination you take their contact details and ask where they live so that you are able to trace the person in case their phone is off, and you call community worker of that area…they sometimes give us numbers that are not working and they go straight to voicemail. And then now you must look for a community worker for that place so now it becomes a long process.” Nurse, Primary, Eastern Cape*

> *“If we don’t get hold of the clients… It tells you to send CHWs… They bring the client to us.” Nurse, Primary, Eastern Cape*

> *“A few weeks ago, we went to visit [area] to find out what services they offer. …So, what we have been doing, for example, for patients where we worry that there’s a cancer, we’ve taken a biopsy. We used to phone, phone, phone, we can’t get hold of the patient. We phone the police … I would never have bothered to send police because it’s quite scary to send police to a patient’s home when they are community workers whose main job, and I imagine the more patients that they are able to track, then they get the salary.” Doctor, Western Cape, Tertiary*

In both countries (but particularly notable for Zimbabwe and the Eastern Cape, SA), HCWs in both the in-depth interviews and clinical advisory workshops perceived that patients’ preferences for traditional healers could result in delays in attending referral appointments. Particularly pertinent for Zimbabwe, HCWs reported that some patients believe that cancer has a spiritual cause and therefore needs traditional intervention.

> *“Timely referral is when someone has a problem and comes to you immediately. People try elsewhere. We are Africans and we would try what our ancestors or what great and whatever used to do, and we dabble that thereby reducing our chances of treating early.” Nurse, Tertiary, Bulawayo*

> *“Some believe in going to traditional healers, some believe in the apostles. Most of them after they go they will realise that ah, like when it comes to cancer, I give examples that with cancer […], they would take a long time while they were bleeding and they would say that it will end, and they will be going for prayers.” Nurse, Primary, Harare*

### Healthcare provider and system factors

#### Poor referral and feedback systems

PHC practitioners from both countries in the interviews and clinical advisory workshops reported challenges related to the processes for referral to secondary and tertiary facilities, including not being able to contact the referral department timeously to provide patients with a date for their appointment while they are with them at PHC. As a result, HCWs need to contact patients again, which is complicated by patients not being contactable by phone or updating their address. This means that referral is delayed, and some patients are lost to follow-up. Additionally, PHC practitioners in the clinical advisory groups in SA reported that referral processes within facilities differ according to staff role, with some staff members not able to access computers to do referrals.

> *“In order to refer to the SOPD [Surgical outpatient department], we have to send them an email. And often, we’re waiting weeks for an email reply. Then they reply to us, and then they say, here’s the date for the patient, please inform the patient. Now this is weeks later, and then you try and phone the patient on the number that they’ve given.” Doctor, Primary, Western Cape*

PHC practitioners state that they do not receive feedback about referred patients, so they remain unsure if the referral was appropriate, what the patient’s diagnosis was or what continuous care is needed at a PHC level. Additionally, in the clinical advisory workshops, PHC practitioners reported a general lack of awareness of referral pathways and services available at each facility. One participant in the Eastern Cape, SA reported refusing to do the repeat Pap smear on a patient who had been sent back from secondary to primary without clear reason. In both the in-depth interviews and the clinical advisory workshops, HCWs perceived that more communication and feedback to PHC level would improve referral, strengthen PHC practitioner’s knowledge of cancer symptoms and inform PHC practitioners whether their referral was appropriate.

> *“…when we get feedback, then maybe we know that when we referred this patient, did they get the help that they needed? What was done for the patient and how long did it take? Did the patient achieve anything from that referral?” Nurse, Primary, Bulawayo*

> *“…sometimes you’ll receive results that say refer for colposcopy and then they would send the person back and say they must do examination again. And I would ask myself why they would refer for colposcopy in the first place only to be sent back to the clinic.” Nurse, Primary, Eastern Cape*

One notable exception that was mentioned in both in-depth interviews and the clinical advisory workshops related to referral systems that used an online referral platform called JotForm to book appointments immediately and has an inbuilt healthcare practitioner education component. This is related specifically to breast cancer in the Western Cape, SA. Better communication is also facilitated by the network of primary, secondary and private GPs that was set-up by the clinical team in that department. Through this platform, PHC HWCs are able to immediately book secondary care appointments and provide dates during consultations, which reduces loss to follow-up and referral delay. Additionally, referring HCWs are provided with real-time feedback on their referral by selecting patient symptoms in-app and getting immediate information on whether the referral is appropriate. Further details on HCWs’ experience of using these digital health tools are discussed in Arendse *et al*.^44^

> *“And if we do feel anything suspicious, we then refer online to [Tertiary Hospital]. And they then give us… We can choose an appointment date and time. They’re quite flexible, and they are very accommodating with their dates because you can go for the next day if they have availability.” Nurse, Primary, Western Cape*

#### Training needs

HCWs in both in-depth interviews and clinical advisory workshops reported that they had training needs related to symptom appraisal and management of cancer symptoms, and risk factors – particularly for colorectal cancer. The HCWs in the clinical advisory group noted that it was not enough to only know what symptoms of cancer, but also know what to do next. Due to a focus on HIV at a PHC level in both countries, PHC practitioners perceived that they generally had better knowledge of cervical cancer symptoms and risk factors yet HCWs at a tertiary level in the Eastern Cape reported that newly trained nurses and medical officers do not receive sufficient practical training to do pelvic examinations, Pap smears and punch biopsies.

> *“…there are learning gaps as some of our nurses are not trained [in managing patients with possible breast, cervical or colorectal cancer symptoms].” Nurse Manager, Primary, Bulawayo*

> *“Yes, there is definitely gross lack of training. Gross lack of training from the training institutions that train. That do the training. For instance, this thing for the nurses, many nurses come out of nursing school without knowing how to actually examine the pelvis, let alone take a Pap smear. And, for the medical students, for the medical training, medical students come out of medical school, despite the high prevalence of cervical cancer in their training institution, they have never examined a patient with cervical cancer. Let alone, taken a punch biopsy. So, who’s going to teach them?” Doctor, Tertiary, Eastern Cape*

In both countries, HCWs reported that training should not be a one-off, but lifetime learning as healthcare advances. Currently, HCWs reported that further professional development is lacking, and information is imparted through other colleagues and acquired institutionally. Particularly in the Eastern Cape, SA, primary HCWs reported learning how to do procedures, such as pelvic examinations and Pap smears, from other colleagues during routine practice. In Zimbabwe, HCWs noted that many senior HCWs have left the sector, resulting in young nurses and doctors being the predominant make-up of the workforce and that they need sufficient training to make up for this loss of senior professionals.

> *“We all need to be taken to a thorough training and a proper training. Look, when I started doing Pap smears, we were using spatulas and those lights before. And it’s not like even then I was trained. It was an information imparted to me by my colleague.” Nurse, Primary, Eastern Cape*

> *“We have got many of our senior health workers are out of the country, brain drain. So, we have got young nurses, young clinicians who are just coming from school. I think if they are empowered it will be easy. It will save our community because they will quickly diagnose-, they will quickly know the signs and symptoms of cancer so that when the patient presents to them, they are transferred there and there… [They should be trained on] the cause of cancer, possible causes of cancer, history taking and how to examine. I think that those are the key.” Nurse, Primary, Harare*

#### Awareness of protocols and guidelines

PHC practitioners across both countries reported that they were not aware of formal protocols and guidelines for managing and referring patients with suspected cancer symptoms. The referral procedures used are often informal, reportedly not written up and are held by institutional knowledge. Occasionally, this results in “incorrect” referrals, for example, referring to the wrong department. Internal procedures and guidelines are not standardised nor uniformly available at facility level. Resultantly, HCWs rely on knowledge from national guidelines, training, and institutional knowledge.

> *“I’m not aware of any formal protocol, no.” Doctor, Primary, Western Cape*

> *“I haven’t seen [protocols and guidelines] now.” Nurse, Primary, Eastern Cape*

> *“Ah, no we don’t have the protocols.” Nurse, Primary, Harare*

> *“Actually, perhaps I am the only ignorant person about the guideline. The guideline is there but I think its emphasis on and retraining of cadres because you get new people because of the brain drain.” Nurse, Tertiary, Bulawayo*

#### Inappropriate and suboptimal clinical assessments

Referral pathways for patients with suspected cancer symptoms are impacted by inappropriate and suboptimal clinical assessments. Secondary and tertiary HCWs in both in-depth interviews and clinical advisory workshops reported that they see patients who have had multiple visits to PHC prior to referral where the patient has received inappropriate management, such as antibiotics for an extended period, Pap smears when they have visible cervical lesions, or repeated treatment of rectal bleeding as haemorrhoids without further investigation. This mismanagement is reported to result in significant diagnostic delays.

> *“Once that patient communicates that to a doctor who is not going to examine and confirm, and that doctor will just write only on the file, it means it’s already labelled on the file. Every doctor that will see that patient, will think haemorrhoids, haemorrhoids. No one bothers to examine the patient. And you just give this treatment for constipation and haemorrhoids.” Doctor, Secondary, Eastern Cape*

> *“…sometimes when a patient goes be it to a secondary or tertiary facility, for instance, if a patient presents with vaginal discharge or post coital bleeding, at times the patient is mismanaged, like she will be treated for sexually transmitted infections. The patient is given some antibiotics, maybe for 7 days or be it 2 weeks. The patient comes back, the discharge has not resolved, and she is given some antibiotics again yet there is disease progression.” Nurse, Tertiary, Harare*

> *“And then if they get HCWs who have got no experience when it comes to tumours, the HCW will see the patient and send the patient home. I’ve seen cases like that. They refer the patient home and say, if [the breast lump] doesn’t give you pain, it’s not bothering you, don’t stress.” Doctor, Secondary, Eastern Cape*

PHC practitioners reported that asymptomatic patients are diagnosed through regular Pap smears, and that it is very rare to find a patient with symptoms of cervical cancer. However, doctors working in tertiary care in SA and Zimbabwe in both the in-depth interviews and clinical advisory workshops stated that cervical cancer patients are still arriving with advanced stage disease and inappropriate or irregular Pap smears have been done at PHC on symptomatic patients. Participants in SA reported this to have resulted in delays in referral by up to eight months. Tertiary HCWs in the clinical advisory workshops reported that vaginal discharge syndrome is common misdiagnosis among women with symptomatic cervical cancer. HCWs in secondary and tertiary care perceived that PHC practitioners do not visualise the cervix when patients’ present with vaginal discharge and prescribe antibiotics repeatedly over an extended period. They state that this is due to insufficient PHC practitioners’ education, overworked healthcare system, PHC practitioners’ reluctance to do vaginal examinations, and lack of available speculums.

> *“And you phoned that sister in charge of the clinics… this patient that I’ve got in front of me has had antibiotics prescribed for the past nine months. Not a pelvic examination. She is a patient who’s been attending your clinic, HIV positive, for the past so many, and has never had a Pap smear.” Doctor, Tertiary, Eastern Cape*

While a few HCWs could name colorectal symptoms, many PHC practitioners reported never managing a patient they suspected of colorectal cancer or HCWs reported that digital rectal examinations are not done at primary care level for patients with colorectal symptoms. Furthermore, a secondary level HCW in the Western Cape reported that colorectal symptoms are vague, and patients represent multiple times at a PHC level.

> *“And then I find the patients will often go to the clinic doctor, and those doctors, or nurse or whomever they see, are so busy. And I think because colorectal symptoms are quite subtle and they’re quite common, my experience is that patients re-present multiple times often to the clinic before they finally achieve a referral.” Doctor, Secondary, Western Cape*

#### Resource limitations

Resource constraints were a major limitation widely reported by participants in both and clinical advisory workshops, including consumables, theatre time, beds, human resources, and specific infrastructure (such as endoscopy, mammogram, computerized tomography (CT) scan, and magnetic resonance imaging). Clinicians reported long waiting lists and broken machines that hamper timely access to diagnosis. For example, some facilities do not have access to certain diagnostics or machines are not functioning with no plans for repair.

> *“A challenge almost arises with the tools. Because we’ll order the buffers, the brushes, and we won’t get them.” Nurse, Primary, Eastern Cape*

Mentioned in both in-depth interviews and clinical advisory workshops, insufficient health facility provided patient transport infrastructure within health facilities in SA reportedly causes delays for patients having to travel to secondary and tertiary health facilities for further diagnostics, investigations, and treatment. Tertiary facilities serve large geographic areas, requiring patients in rural areas to travel long distances to seek care. Across both provinces, transport availability in rural areas is limited, requiring HCWs to book patients’ appointments in accordance with transport availability, which can result in diagnostic delays.

> *“So, there’s a planned patient transport bus … But that’s a limited number of seats… But it can happen that patients miss their dates, because something goes wrong with the transport.” Doctor, Primary, Western Cape*

> *“… the transport issue. Now patient is discussed, got that particular date, and when the patient goes to book the transport, there’s no transport. They end up missing the date. You have to rebook them. They get a date that is far then finding that the year is finishing, six months is finishing. Patient has been in the system but does not get to the tertiary institution. And there is still going to need to be assessed, biopsied, wait for the results and referred to.” Doctor, Secondary, Eastern Cape*

## Discussion

Improving timely diagnosis of breast, cervical and colorectal cancer in Southern Africa requires strategies and interventions that are responsive to both patient-related and health system delays and challenges in the pathway from symptoms to diagnosis.^6, 12, 13^ This study examined patient-, HCW-, and health system factors that impacted the Diagnostic interval. In line with other studies, this study found that patient-level delays in the diagnostic interval were due to lack of access to transportation, rurality and distance to healthcare facilities, preference for alternative medicines and availability and affordability of medical care.^27^

Particularly for Zimbabwe, the financial cost of healthcare within a socioeconomically deprived context results in substantial delays throughout the care continuum. The economic crisis in Zimbabwe, which started in the 1990s, has resulted in the deterioration of health infrastructure, which is articulated by public health facility closures, lack of medical supplies, insufficient healthcare workers, and lack of financial resources for the maintenance and upgrading of health infrastructure.^48, 49^ Because of cuts in renumeration and allowances, Zimbabwe is experiencing a loss of HCWs to the private sector and immigration which placed additional burden on those remaining.^49^ Embedded within this broader context, there is a significant lack of financial allocation for cancer services.^50^ HCWs and patients accessing cervical and breast cancer services in Zimbabwe reported that barriers to cancer care included insufficient human resources, limited training for HCWs, too few cancer specialists, medicine and equipment shortages, reliance on out-of-pocket payments by patients, and lack of clear referral systems.^50–52^ Furthermore, centralised services make it difficult for rural patients to access health facilities.^50^ Considering these challenges, interventions aimed at improving the diagnostic interval must be tailored to the context-specific socio-economic realities of health systems.

Decentralising diagnostic services from tertiary to primary institutions^53–55^ and patient navigation^5, 55–58^ have proven to be promising interventions for structural barriers affecting referral pathways by prompting patients to move along the referral pathway, improving access to health facilities and reducing the burden on tertiary-level facilities. Evidence from Kenya, Botswana and South Africa demonstrated that decentralising diagnostic services can reduce the workload of tertiary hospitals, and limit the burden on patients to travelling far distance to access healthcare services, and improved diagnostic turnaround time.^53–55, 59^ Challenges associated with decentralisation of healthcare in LMICs included inadequate technical skills needed to perform tasks, insufficient resources for essential services, and decentralisation of decision-making without granting authority to implement decisions.^60^ A systematic review of decentralisation in LMICs found that decentralization led to a deterioration of human resources, medicines, and equipment, primarily due to increased bureaucracy and insufficient managerial skills at the local level.^60^ To overcome these barriers, community participation and engagement in health planning processes are essential to improve communication and accountability between communities and the health sector.^60^

Patients experience fragmented care from initial symptom experienced through the entire care journey.^61^ Studies conducted in LMICs – primarily conducted in the African region and South Asia - have shown that patient navigation for early detection of breast cancer, with and without mHealth, has been successfully used to improve timely diagnosis.^5, 56–58, 62^ These studies have shown that patient navigators and CHWs play a crucial role in doing initial clinical breast examinations, making onward referrals, facilitating access to specialist care services and connecting healthcare providers with patients by tracking those who have missed their appointments.^5, 56–58, 62^ However, training for patient navigators is variable across contexts.^62^ Within South Africa and Zimbabwe, CHWs are a recognised and important category of HCWs – linking the community with PHC facilities – who could potentially assist with navigation of symptomatic patients. Incorporating an evidence-based, contextually sensitive patient navigation programme into CHW training may be a feasible intervention to improve patient referral.^58^ However, in South Africa and Zimbabwe, CHWs are employed by community-based organisations and face challenges such as limited resources, precarious employment, low or no pay, poor role clarity, and insufficient logistical and health systems support.^63–65^ The CHW space is predominantly donor driven, which adds precarity to their position within the sector.^63, 65^ Within the cancer care continuum, patient navigators in South Africa are currently used in the research and NGO space predominantly for post diagnosis navigation.^66–68^ These services have yet to be translated into navigation for symptomatic patients pre-diagnosis and are also limited by external sources of funding.^66^

A study in Bangladesh demonstrated that mHealth tools in conjunction with patient navigation from symptom detection at PHC to diagnosis allowed for improved reach, provided better data quality, and encouraged more patients with breast symptoms to attend further care.^5^ mHealth is the use of mobile devices, such as smart phones and wearable devices, to support and improve healthcare. However, the findings of our study suggest that the use of mHealth may be complicated by patients not being contactable via phone and HCWs requiring to use their own electronic devices for clinical work (see Arendse et al.^44^ for an extended discussion). However, there are examples of successful mHealth interventions in other health areas, such as HIV, sexual and reproductive health, noncommunicable diseases and maternal care, in South Africa and Zimbabwe.^69–74^ Designing and implementing successful mHealth interventions to improve earlier diagnosis of cancer in Southern Africa must draw on lessons from previous studies, and take participatory and human centred design approaches which allow for the creation of toolkits that are contextually relevant and accessible.^71, 75^ For example, choosing text-based as opposed to a smart phone app based approach may be more accessible.^69, 71^ mHealth toolkit development to improve earlier diagnosis of cancer needs to build in flexibility to cater to context-specific technology realities and needs.

Several studies in Southern Africa have indicated that patients often visit PHC facilities multiple times before being referred.^12, 26, 76^ In this study delays in referral from PHC were attributed to misappraisal and mismanagement of possible cancer symptoms at PHC level and insufficient training for PHC practitioners. Cancer symptoms often overlap with common conditions, complicating timely referral.^26^ This study identified a need for continuous HCW training and professional development to improve cancer symptom assessment, particularly at PHC level. Training interventions in studies of various cancer types in LMICs aimed at enhancing the clinical skills of healthcare professionals have demonstrated consistent improvements to time to diagnosis.^55, 77^ Intervention studies in LMICs focused on enhancing the diagnostic capacity of HCWs, utilising methods such as video training, audio-visual education material, discussions, presentations, roleplaying and journal reviews, demonstrated improved knowledge scores for healthcare professionals.^55, 78–84^ Interventions designed to enhance diagnostic knowledge and skills have been shown to reduce diagnostic turnaround time,^55, 59^ increase detection rates,^18, 55^ and promote downstaging.^18, 19, 55, 85^ However, interventions to improve diagnostic knowledge and skills cannot be done as once offs as HCWs require regular training.^79^ While standardised training is imperative for bridging gaps in education to improve HCW cancer symptom assessment and appraisal, developing interventions that build in continuing education on cancer symptoms and immediate feedback into everyday clinical practice, such as referral tools, can be a cost-effective and targeted solution for current knowledge gaps.

Scholarship in similar contexts has shown that a lack of standardised referral pathways negatively impacts the diagnostic interval by causing unnecessary patient delays.^21, 23, 27^ The lack of a uniform approach can result in unequal access to care, delays for patients, inappropriate referrals, and an increased burden on primary HCWs who must keep track of multiple referral pathways.^21, 23, 27^ In this study, referral pathways varied within and between health facilities and were often held in institutional knowledge rather than by using standardised protocols. Feedback on referrals back to PHC providers are not common practice, hindering opportunities for continuing education for PHC practitioners.^26^ Poor referral pathways are underpinned by poor communication practices within and between facilities. The predominant format of referral pathways results in long appointment waiting times and patients being lost to follow-up. Technological interventions for patient referrals, follow-ups, and clinical decision-making have proven feasible even in rural settings, provided that the necessary infrastructure and operational readiness are in place.^5, 22, 55, 86^ Developing locally specific and effective interventions to improve referral pathways should be underpinned by current best practices, patient realities and health system limitations.

To our knowledge, this is the first study in SA and Zimbabwe to investigate HCWs’ across different levels of care perceptions of the challenges and facilitators in the pathways to diagnosing breast, cervical, and colorectal cancers. The qualitative nature of the inquiry and strong theoretical underpinning with the Model of Pathways to Treatment enabled an in-depth exploration of the research question. Conducted in both countries, the study examined perceived variations in healthcare systems across Southern Africa. By examining cancer care in two distinct countries, the study provides valuable insights into how different levels of human development index impact healthcare systems and identify unique challenges and best practices. A comparative analysis reveals important differences in health outcomes and resource allocation, which can inform targeted interventions. This study also was conducted in both urban and rural settings to explore geographic differences in service provision, offering an examination of similarities and differences across geographical contexts. A limitation of this study is that it provides HCW perceptions of patient-level factors, which may not entirely reflect patient experiences, beliefs and circumstances.

## Conclusion

Improving the diagnostic interval for breast, cervical, and colorectal cancer in Southern Africa necessitates targeted strategies that address both patient- and health-system related factors with consideration given to socio-cultural and economic contexts.

## Data Availability

The data related to the preparation of this study can be made available on request.

## Abbreviation List

ART: Antiretroviral therapy
AWACAN-ED: African aWAreness of CANcer & Early Diagnosis programme
CHW: Community Healthcare Worker
eHealth: Electronic health
GP: General Practitioner
HCW: Healthcare worker
HDI: Human Development Index
HIV: Human immunodeficiency virus
IQR: Interquartile range
LEEP: Loop electrosurgical excision procedure
LMIC: Low-and-middle-income country
mHealth: Mobile health
OI: Opportunistic infection
OPD: Outpatient department
PHC: Primary healthcare
SA: South Africa
VIAC: Visual Inspection with Acetic Acid and Cervicography
WHO: World Health Organization

## Authors contributions

SD was responsible for project management in South Africa; data collection in South Africa; data analysis; and leading the write-up of the manuscript. SM and MM were responsible for data collection in Zimbabwe; data analysis; and reviewing the manuscript for critically for important intellectual content. KDA and NT were data analysis; and reviewing the manuscript for critically for important intellectual content. SES and TR was responsible for reviewing the manuscript for critically for important intellectual content. VAS was responsible for the data analysis and reviewing the manuscript for critically for important intellectual content. BTG was responsible for project management in Zimbabwe and reviewing the manuscript for critically for important intellectual content. FMW and JM carry over-all end-responsibility of the project, including oversight for the protocol development, implementation, and assurance for the timely reporting and dissemination of study results.

## Data sharing statement

The data related to the preparation of this study can be made available on request.

## Patient Public Involvement

Public patient involvement is central to the AWCAN-ED Project. The AWACAN-ED steering committee and broader collaborator team, which includes HCWs, policy makers and patients in the South African and Zimbabwean healthcare system, have been involved from the onset in the development of the initial funding application, proposal development, data collection and provided feedback on initial findings during feedback workshops.

