## Supplemental Files for "Challenges and facilitators in pathways to cancer diagnosis in Southern Africa: A qualitative study"

**Supplementary file 1a: Primary HCW In-Depth Interview Guide**

1. Welcome & introduction

- Introduce self, role,
- Introduction to research: In this project we are interested in your views about diagnosing and managing patients with breast, cervical and colorectal cancer and your views on new ways to encourage prompt diagnosis and treatment.
- Hand out information sheet and go through it with participant.
- Discuss confidentiality and anonymity.
- Use of audio-recording and data storage
- Length of interview and nature of discussion (specific topics to cover but will be like a conversation, ‘there are no right or wrong answers’)
- Any questions?
- Consent (confirm understanding of study and consent participant)
- Before we start with the main interview can I ask for some details about yourself so we can have a record of the people we interviewed?
  - Hospital name and location
  - Role / job title and site
  - Years in role
  - Gender
  - Age
  - Highest Education level
- *Begin audio-recording*

2. Current practice*^[1]^*

- To start off, can you tell me about your role in the facility? What are your main duties? *Clarify if patient facing role*
- Prompt: We are interested in your views about diagnosing and managing patients with possible breast, cervical and colorectal cancer. Which of these types of cancer are involved in your role?

*2a. Management of patients with symptoms*

- Can you talk through the typical journey of a patient who presents with possible symptoms of *[breast/cervical/colorectal]* cancer)?
- Prompt: Please talk about the processes and people involved, what happens when a patient arrives and whom do they see first?
- Are there any challenges for managing these patients?
- For participants with patient-facing roles:
- What symptoms/factors would spark a high suspicion of cancer?
- What would make you less inclined to suspect cancer?
- What do you use to support your decision making?
- In your current practice, is there a recommended protocol for managing patients with possible symptoms of *[breast/cervical/colorectal]* cancer?

*2b. Investigations*

- If you suspect [breast/cervical/colorectal] cancer, are you able to do any tests to confirm or rule out this suspected diagnosis in this facility?
- In the current system, what helps prompt/timely investigations/tests when *[breast/cervical/colorectal]* cancer is suspected? What works well at present?
- In the current system, are there problems with getting prompt/timely investigations when *[breast/cervical/colorectal]* cancer is suspected? If yes, where do things go wrong at present? [Prompts: challenges faced, what could be improved?]

*2c. Referral*

- How does the referral system work for patients with possible or suspected *[breast/cervical/colorectal]* cancer?
- What helps prompt/timely referral to a secondary/tertiary healthcare facility when *[breast/cervical/colorectal]* cancer is suspected?
  - (Prompt: What works well at present?)
- In the current system, what problems are there with referral to a secondary/tertiary healthcare facility when *[breast/cervical/colorectal]* cancer is suspected?
  - (Prompt: Where do things go wrong at present?) [Prompts: challenges faced, what could be improved?]

*2d. Diagnosis*

- In the current system, what do you think helps prompt/timely diagnosis of *[breast/cervical/colorectal]* cancer at secondary/tertiary facilities?
  - (Prompt: What works well at present?)
- In the current system, what problems do you think there are with getting prompt cancer diagnosis once someone attends secondary/tertiary facilities?
  - (Prompt: Where do things go wrong at present?)
- [Prompts: challenges faced, what could be improved?]

*2e. What would help?*

- What do you think would help to improve prompt diagnosis of cancer? Use the below ideas for prompts:
- Guidelines for who to refer or when / where to refer?
- Training of/ education for healthcare professionals [to consider cancer]?
- What needs to be covered?
- How should training be delivered? (Online? How comfortable are you with using computers / mobile applications for professional educational purposes?)
- Fast track referrals for patients with suspected cancer?
- Electronic booking appointments?
- Feedback about referrals?
- Is there anything else that you think may help?

Technology Acceptance and Usability

We are interested in your experiences of using technology in the workplace as we are exploring if this could help in the context of cancer diagnosis. In the next section we will ask you questions about your experiences.

*3a Communicating clinical information*

- What is the main way you communicate clinical information with your colleagues?
- Does your facility use an Electronic Health Record (EHR) System?
- If yes, can you tell me more about your views of the system(s) (include names)
- Can you tell me about any challenges of using the EHR system?
- Can you tell me about any the benefits of using the EHR?
- Do you use the EHR System for patients with possible cancer symptoms, if so in what ways? (i.e. for referral)

*3b Technology supported practice*

- What is your experience using computers or mobile applications for clinical purposes?
- Prompt: Can you tell me about what you have used, what has worked well?
- Prompt: What are the challenges or barriers in using computers or mobile applications for clinical purposes? Do these devices support clinical decision making?
- Prompt: Would the use of a computer or mobile health application fit well within your clinical practice for managing patients who present with possible symptoms of cancer?
- Can you say why/why not?
- Are there any staff training needs related to the use of technology for clinical practice? Ask for detail.
- Do you think staff within the health facility are motivated to use computer or mobile phone-based tools to support clinical practice?
- Are there certain patients/patient groups for whom online or mobile phone-based tools would be more/less suitable?

*3c Technology provision*

- Are you provided with any devices (computer, smart phone, basic phone, tablet, laptop) to support clinical practice by this health facility? This may include shared devices. (Ask for details)
- Prompt: Do you use any personally owned technological devices to support clinical practice?
- Prompt: Who provides funding for your access to the internet/Wi-Fi for your clinical practice?
- If personal, can you tell me more about how you go about accessing the internet and whether internet access is affordable for you?

*3d Technological infrastructure*

- If/when the health facility loses electricity, how does it impact your work?
- Have you made any changes in your practice to accommodate for periods when electricity goes out? (If yes, ask for details)
- If/when you don’t have access to the internet/Wi-Fi, how does it impact your work?
- When a device malfunctions at your hospital how long does it take to get a repair?
- Is there an ICT (computing support) team that repairs technological devices at your facility?

Closing

- Thank you very much – that’s all my questions. Is there anything else you would like to add?
- We are speaking to a range of facility managers and healthcare professionals. Is there anyone else who you think may be useful to interview? If so, ask if willing to forward study invitation to them.
- Thank participant and close interview (confirm consent and confidentiality) and confirm that they would be interested in a copy of the final report/ write up of the study.

**Supplementary file 1b: Secondary/Tertiary Care Interview Guide**

1. Welcome & introduction

- Introduce self, role, Introduction to research: In this project we are interested in your views about diagnosing and managing patients with breast, cervical and colorectal cancer and your views on new ways to encourage prompt diagnosis and treatment.
- Hand out information sheet and go through it with participant.
- Discuss confidentiality and anonymity.
- Use of audio-recording and data storage
- Length of interview and nature of discussion (specific topics to cover but will be like a conversation, ‘there are no right or wrong answers’)
- Any questions?
- Consent (confirm understanding of study and consent participant)
- Before we start with the main interview can I ask for some details about yourself so we can have a record of the people we interviewed?
  - Hospital name and location
  - Role / job title and site
  - Years in role
  - Gender
  - Age
  - Highest Education level
- *Begin audio-recording*

2. Current practice*^[2]^*

- To start off, can you tell me about your role in the facility? What are your main duties? *Clarify if patient facing role*
- Prompt: We are interested in your views about diagnosing and managing patients with possible breast, cervical and colorectal cancer. Which of these types of cancer are involved in your role?

*2a. Management of patients with symptoms*

- Can you talk through the typical journey of a patient who presents with possible symptoms of *[breast/cervical/colorectal]* cancer)?
- Prompt: Please talk about the processes and people involved, what happens when a patient arrives and whom do they see first?
- Are there any challenges for managing these patients?
- For participants with patient-facing roles:
- What symptoms/factors would spark a high suspicion of cancer?
- What would make you less inclined to suspect cancer?
- What do you use to support your decision making?
- In your current practice, is there a recommended protocol for managing patients with possible symptoms of *[breast/cervical/colorectal]* cancer?

*2b. Investigations*

- If you suspect [breast/cervical/colorectal] cancer, are you able to do any tests to confirm or rule out this suspected diagnosis in this facility?
- In the current system, what helps prompt/timely investigations/tests when *[breast/cervical/colorectal]* cancer is suspected? What works well at present?
- In the current system, are there problems with getting prompt/timely investigations when *[breast/cervical/colorectal]* cancer is suspected? If yes, where do things go wrong at present?
  - [Prompts: challenges faced, what could be improved?]

*2c. Referral*

- How does the referral system work for patients with possible or suspected *[breast/cervical/colorectal]* cancer?
  - (Prompt: Are patients referred here from other facilities and do you refer patients elsewhere?)
- What helps prompt/timely management of patients who are referred to this facility when *[breast/cervical/colorectal]* cancer is suspected?
  - (Prompt: What works well at present?)
- In the current system, what problems are there managing patients referred to this facility when *[breast/cervical/colorectal]* cancer is suspected?
  - (Prompt: Where do things go wrong at present?) [Prompts: challenges faced, what could be improved?]

*2d. Diagnosis*

- In the current system, what helps prompt/timely diagnosis of *[breast/cervical/colorectal]* cancer at secondary/tertiary facilities?
  - (Prompt: What works well at present?)
- In the current system, what problems are there with getting prompt cancer diagnosis once someone attends secondary/tertiary facilities?
  - (Prompt: Where do things go wrong at present?)[Prompts: challenges faced, what could be improved?]

*2e. What would help?*

- What do you think would help to improve prompt diagnosis of cancer? Use the below ideas for prompts:
- Guidelines for who to refer or when / where to refer?
- Training of/ education for healthcare professionals [to consider cancer]?
- What needs to be covered?
- How should training be delivered? (Online? How comfortable are you with using computers / mobile applications for professional educational purposes?)
- Fast track referrals for patients with suspected cancer?
- Electronic booking appointments?
- Feedback about referrals?
- Is there anything else that you think may help?

Technology Acceptance and Usability

We are interested in your experiences of using technology in the workplace as we are exploring if this could help in the context of cancer diagnosis. In the next section we will ask you questions about your experiences.

*3a Communicating clinical information*

- What is the main way you communicate clinical information with your colleagues?
- Does your facility use an Electronic Health Record (EHR) System?
- If yes, can you tell me more about your views of the system(s) (include names)
- Can you tell me about any challenges of using the EHR system?
- Can you tell me about any the benefits of using the EHR?
- Do you use the EHR System for patients with possible cancer symptoms, if so in what ways? (i.e. for referral)

*3b Technology supported practice*

- What is your experience using computers or mobile applications for clinical purposes?
- Prompt: Can you tell me about what you have used, what has worked well?
- Prompt: What are the challenges or barriers in using computers or mobile applications for clinical purposes? Do these devices support clinical decision making?
- Prompt: Would the use of a computer or mobile health application fit well within your clinical practice for managing patients who present with possible symptoms of cancer?
- Can you say why/why not?
- Are there any staff training needs related to the use of technology for clinical practice? Ask for detail.
- Do you think staff within the health facility are motivated to use computer or mobile phone-based tools to support clinical practice?
- Are there certain patients/patient groups for whom online or mobile phone-based tools would be more/less suitable?

*3c Technology provision*

- Are you provided with any devices, (computer, smart phone, basic phone, tablet, laptop) to support clinical practice by this health facility? This may include shared devices. (Ask for details)
- Prompt: Do you use any personally owned technological devices to support clinical practice?
- Prompt: Who provides funding for your access to the internet/Wi-Fi for your clinical practice?
- If personal, can you tell me more about how you go about accessing the internet and whether internet access is affordable for you?

*3d Technological infrastructure*

- If/when the health facility loses electricity, how does it impact your work?
- Have you made any changes in your practice to accommodate for periods when electricity goes out? (If yes, ask for details)
- If/when you don’t have access to the internet/Wi-Fi, how does it impact your work?
- When a device malfunctions at your hospital how long does it take to get a repair?
- Is there an ICT (computing support) team that repairs technological devices at your facility?

Closing

- Thank you very much – that’s all my questions. Is there anything else you would like to add?
- We are speaking to a range of facility managers and healthcare professionals. Is there anyone else who you think may be useful to interview? If so, ask if willing to forward study invitation to them.
- Thank participant and close interview (confirm consent and confidentiality) and confirm that they would be interested in a copy of the final report/ write up of the study.

**Supplementary File 2: Clinical Advisory Workshops**

*Table 3: Clinical advisory group workshop overview*

| **Clinician Workshops** | **South Africa** | | **Zimbabwe** | |
| --- | --- | --- | --- | --- |
| **Date** | 18-Mar | 23-May | 18-Mar | 26-Mar |
| **Location** | Cape town | Cape Town | Harare | Bulawayo |
| **Number of clinicians** | 6 | 6 | 7 | 7 |
| **Clinician summary** | PHC & Specialists | Specialists | 3 PHC, 4 Specialists | 3 PHC, 4 Specialists |
| **Cancers covered** | Breast & cervical | Colorectal | Breast, Cervical & Colorectal | Breast, Cervical & Colorectal |
| **Clinician roles** | Family physicians Endocrine and Breast Surgeon Gynaecological Oncologist Gynaecologist | Colorectal surgeons Registrars Oncologist | Gynaecological Oncologist Surgical Gastro-oncologist General Surgeon (Breast) Medical Officer Clinical Medical Officer GMO Clinical Oncologist | Gynaecologist Surgeon, Breast Specialist General Surgeon, Colorectal Specialist General Practitioner Clinical Medical Officer GMO Oncologist |

Table 4: Summary of themes emerging from clinical advisory workshops

| **Themes** | **Summary** |
| --- | --- |
| HW Training & education | The nurses as the first point of contact were seen as key and they have a wide variety of training and experience.  Across cancers it was said that training was needed on recognising symptoms and knowing what to do next - for all healthcare workers  Primary care reports of not being aware of referral pathways and general lack of knowledge of services available - so better marketing and information needed.  Particular concerns about the value of current clinical breast examinations and breast health policy not being implemented in SA. |
| Feedback to Primary Healthcare | General feeling that more feedback and communication back to primary is needed and would help improve matters.  Primary care keen to have feedback on whether referrals were appropriate - seems to happen in Gynae in SA now. |
| System/Approach changes to improve pathway | Better use of existing resources and more resource for cancer is needed. System is overburdened currently Communication issues and lack of electronic data capture discussed as issues.  An improved referral pathway through the system generally felt will reduce delays and improve matters.  Biopsies mentioned with differing views - SA view to centralise biopsies to speed up process, Zim wanted decentralisation to reach more people and training/upskilling on how to do biopsy.  Difference reported between local & rural patients - seen as a failure of the Health System. |
| Patient fear, stigma & beliefs | Patients believe cancer is a death sentence and fear is cancer is generally acknowledged.  Colorectal cancer patients additionally are afraid of colonoscopy and the surgical procedure and colostomy.  Religious beliefs, belief in alternative medicine, different cultural beliefs, perceived cost of treatment and lack of faith in the system also mentioned as barriers.  Education, counselling services and building trust being the community and health workers seen as the way forward.  *Caveat: these are HCW’s views and assumptions of patient barriers/facilitators to early diagnosis, rather than the perspectives of patients* |
| Costs & Transport | Costs and transport issues mentioned in all workshops as barriers. In Zim costs of scans and tests more of an issue.  Reports of funds running out and people not being able to pay for treatment after diagnosis in Zim.  In SA, reports of people paying for a private diagnosis, then not being able to afford private surgery so come to clinic for treatment.  *Caveat: these are HCW’s views and assumptions of patient barriers/facilitators to early diagnosis, rather than the perspectives of patients* |

1. *For questions 2a-2d:*

   *Repeat questions for breast, cervical and colorectal cancer if their role involves more than one cancer type. Start with the cancer they are most familiar with.*
2. *For questions 2a-2d:*

   *Repeat questions for breast, cervical and colorectal cancer if their role involves more than one cancer type. Start with the cancer they are most familiar with.*
